# Predictors of sex-specific resistance to caTAUstrophe

**DOI:** 10.64898/2026.01.28.26345029

**Authors:** Maria Carrigan, Colin Birkenbihl, Hannah M. Klinger, Oliver Langford, Gillian T. Coughlan, Mabel Seto, Jane A. Brown, Annie Li, Madison Cuppels, Michael Properzi, Jasmeer Chhatwal, Julie Price, Aaron Schultz, Dorene Rentz, Rebecca Amariglio, Harm J. Krugers, Rik Ossenkoppele, Keith Johnson, Reisa Sperling, Timothy J. Hohman, Michael Donohue, Rachel F. Buckley

## Abstract

**INTRODUCTION:** As β-amyloid(Aβ) accumulates, tau pathology spreads beyond medial temporal(MTL) regions into the neocortex, though some older adults resist this progression, or ‘caTAUstrophe’. Given previous evidence of higher tau levels in women, we tested whether tau resistance presents differently in Aβ+ men and women.

**METHODS:** Employing data from 872 Aβ+ older adults(N_Test_=579) across three cohorts, we estimated resistance, defined as the deviation from participant’s expected level of neocortical tau. Models predicting the expected tau levels were trained separately in females(N_Train_=172) and males(N_Train_=121) experiencing ‘typical’ tau progression to assess sex-specific resistance.

**RESULTS:** Relative feature importance in female-only and male-only expectation models differed in 97.7% of variables(p[FDR]<0.001). Moreover, age and Aβ burden predicted male resistance, while CDR, latent PACC, and adjusted hippocampal volume were predictors for both sexes.

**DISCUSSION:** Our study highlights the impact of sex-specific characteristics in the prediction of neocortical tau resistance. Understanding sex-specific resistance pathways informs targeted Alzheimer’s interventions.

## 1. BACKGROUND

Under the ATN framework [1], individuals who are positive for both β-amyloid (Aβ+) and tau (T+) pathology meet the criteria for Alzheimer’s disease (AD) classification, irrespective of cognitive status. Despite the required presence of both pathologies, neocortical (NEO) tau density in particular is linked most strongly to cognitive decline [2,3]. Early tau accumulation in the medial temporal lobe (MTL), in particular the entorhinal cortex, is an early precursor to rapid tau proliferation into the neocortex in preclinical AD [4]. The transition from MTL to NEO tau is not inevitable; while the presence of Aβ and higher MTL tau burden accelerates this ‘caTAUstrophe’ [4], considerable heterogeneity exists in the extent to which an Aβ+ individual might progress to NEO T+. This variability raises an important question: why do some Aβ+ individuals, who may also possess other risk factors including older age, apolipoprotein (*APOE*) ε4 carriership, or female sex, remain low in NEO tau burden (i.e., reflecting NEO tau resistance) while others predictably follow this path? Female sex in particular presents a compelling factor potentially modulating resistance to NEO tau burden, given well-established sex-differences showing higher MTL and NEO tau levels in women[5], faster rates of tau accumulation and proliferation[6], and reduced overall brain resilience to tau pathology[7], compared to men. Therefore, the current study aimed to investigate whether and how resistance to NEO tau burden presents differently in men and women. Among Aβ+ older adults from 3 cohorts, we separately estimated male and female NEO tau resistance using the recently published inverse learning approach[8]. Sex-specific models were built to estimate NEO tau resistance. We first examined how resistance presents in women and men separately (i.e., “what is resistance”) by looking at our expectation models themselves. Subsequently, using our models to estimate resistance in women and men, we tested which factors predict NEO tau resistance in each sex.

## 2. METHODS

### 2.1 Sample characteristics

A total of N=872 Aβ+ individuals across the AD continuum were selected from Anti-Amyloid Treatment in Asymptomatic Alzheimer’s Disease trial and the companion Longitudinal Evaluation of Amyloid Risk and Neurodegeneration study [9] (A4/LEARN, N=388), Harvard Aging Brain Study [10] (HABS, N=72), and Alzheimer’s Disease Neuroimaging Initiative [11] (ADNI, N=412). Across all cohorts, we included participants with demographic data, age, sex, education, *APOE*ε4, as well as longitudinal cognitive performance and longitudinal magnetic resonance imaging (MRI). We conducted this study under the ethical guidelines stipulated by the Partners Human Research Committee, which is the Institutional Review Board for the Massachusetts General Hospital and Brigham and Women’s Hospital. The study was carried out by the guidelines of the Declaration of Helsinki. Written informed consent was obtained from all participants.

### 2.2 Imaging measures

#### MRI

Three-dimensional (3D) structural T1-weighted MRI scans were acquired across all cohorts using previously detailed site-specific scanners [12–14]. T1 images were segmented and parcellated using FreeSurfer software (version 6.0). Hippocampal volume (HV) and total gray matter volume were examined after adjusting for intracranial volume. MRI scans were selected based on closest date to each participant’s tau-PET scan.

#### PET

Tracers for Aβ-PET acquisition were ^18^F-Florbetapir (FBP) in ADNI and A4/LEARN, ^18^F-Florbetaben (FBB) in ADNI, and ^11^C Pittsburgh compound-B (PiB) in HABS. The PET acquisition parameters for each study are described elsewhere [10,15–17]. Distribution volume ratios (DVRs) were calculated using Logan plotting between 40 and 60 minutes post-injection for HABS, and between 50 and 70 minutes post-injection for ADNI and A4/LEARN to develop standardized uptake value ratios (SUVrs). For each cohort, summary measures were derived from a weighted average within a large aggregate cortical region of interest (ROI) that includes the precuneus, rostral anterior cingulate, medial orbitofrontal, superior frontal, rostral middle frontal, inferior parietal, inferior temporal, and middle temporal regions (collectively referred to as FLR). The target composite was referenced to cerebellar grey for HABS, and the whole cerebellum for ADNI and A4/LEARN (FBP and FBB tracers). In ADNI, FBP cortical summary SUVrs were obtained from data previously processed by the University of California Berkeley, accessed through the LONI data portal (http://adni.loni.usc.edu/). In brief, FBP SUVrs were computed by averaging retention values across cortical ROIs from the lateral and medial frontal, anterior and posterior cingulate, lateral parietal, and lateral temporal regions, with reference to the whole cerebellum. As different tracers and processing pipelines were used across cohorts, Aβ-PET signal was dichotomized (positive/negative) using published criteria; 1.11 (FBP) and 1.08 (FBB) SUVr threshold in ADNI [11], 1.15 SUVr threshold and visual read in A4/LEARN [18], and a 1.185 DVR threshold in HABS [19].

For tau-PET, ^18^F-Flortaucipir (FTP) was used in all three cohorts, with study-specific FTP-PET acquisition parameters published previously [20–22]. FTP-PET images were generated based on mean retention between 75 and 105 minutes (ADNI), 90 and 110 minutes (A4/LEARN) and between 80 to 110 minutes (HABS) postinjection. FTP-PET data were processed using in-house pipelines [20], using cerebellar grey as reference region for calculation of SUVrs. Cross-sectional tau-PET composites were calculated for NEO, comprised of bilateral fusiform, middle temporal, inferior parietal, and inferior temporal regions, and MTL, comprised of bilateral amygdala, parahippocampal, and entorhinal regions. Processing of T1-weighted images was performed with FreeSurfer version 6.0 to identify gray-white and pial surfaces, as well as to produce automatic Desikan-Killiany cortical and aseg subcortical ROI parcellations [23]. All quality control measures were carried out as previously described [10].

### 2.3 Cognitive outcome measures

We used the global score of the Clinical Dementia Rating (CDR) [24] and the Preclinical Alzheimer Cognitive Composite score (PACC) [25] for our analyses. Due to different variants of the PACC that are used in each cohort [26], we calculated the latent PACC (lPACC) to harmonize this cognitive composite measure across cohorts. Harmonizing cognitive scores circumvents the drawbacks of standardizing, which imposes strong, and often violated, assumptions on the psychometric properties of the neuropsychological tests [26].

### 2.4 APOE Genotyping

Genotyping of *APOE* was performed in each study through blood sample collection. *APOE*ε4 positivity was determined as heterozygous and homozygous carriers of the ε4 allele (one or two copies, respectively), excluding those carrying the ε2/ε4 combination.

### 2.5 Operationalization of (sex-specific) resistance

Building on previous work[8], we employed an inverse learning approach, a more robust extension of the residual approach, to operationalize NEO tau resistance as a continuous metric. In essence, we select a sample that best portrays what we define as the caTAUstrophe (i.e., low tau resistance as defined by study-specific criteria outlined below). We then use this sample to train an elastic-net regression model that predicts the expected NEO tau-PET burden from a range of predictors (including age, sex, *APOE*ε4, cognition, education, MTL tau SUVr, ICV-adjusted hippocampal volume, and continuous Aβ burden). We call this model the expectation model. This model is subsequently applied to a heterogeneous training-independent sample that has a high likelihood of including individuals with high tau resistance, defined as lower-than-expected NEO tau burden given their characteristics (i.e., the same model predictors as used during model training). The NEO tau resistance estimate is then calculated by subtracting the empirical observed NEO tau burden from the predicted expected tau burden. In this way, resistance is operationalized as the deviation from the predicted expectation (i.e., the error term in a standard residual approach [27]), where a negative residual indicates lower tau resistance while positive residual indicates greater tau resistance. Details on the application of inverse learning to operationalize resistance can be found in the original publication [8]. In this study, the training sample was defined *a priori* following a presumed disease progression according to the ATN framework [1]. As such, we only included individuals with low tau resistance; those who were Aβ+ and T+ at baseline or converted at some point throughout the study to Aβ+ and T+. Tau positivity was defined using a Gaussian Mixture model as explained in the **Supplementary Material**. We included every visit of the selected participants for model training (including any Aβ- and T-visits) to ensure that the model learned to predict expected tau levels across disease stages. We were specifically interested in how the expectation of resistance was estimated in each sex separately, and whether different predictors of resistance became apparent in each sex. Thus, our initial training sample (N=293) was stratified by sex (N_males_=121, N_females_=172), resulting in two separate samples used to train respective expectation models predicting NEO tau burden, one in each sex (male-only/female-only; **Figure1-A**). Demographic information on the male and female training data (expectation sample) is provided in **Supplementary Table** The data used for training the sex-stratified models was not used in any of the further analyses. We estimated sex-specific resistance to NEO tau by applying the female-only expectation model to women (N=316) and the male-only expectation model to men (N=263) separately (**Figure 1-A**). Demographic information is provided in **Tables 1** (Women) and **2** (Men). All linear regression analyses investigating the associations with our predictors of interest were performed on female resistance as estimated in women using the female-only model and male resistance as estimated in men employing the male-only model, respectively (**Figure 1-B**). For the purpose of this study, we will refer to “female-only” and “male-only” as the expectation samples that were used to build the models predicting NEO tau burden in each sex, while “women” and “men” will refer to the heterogenous sample of potentially resistant individuals that each sex-specific model was applied to for estimation of female and male resistance, respectively.

**Table 1.** Female Sample Demographics.

| **Characteristic** | **Overall**<br>**N = 316<sup>1</sup>** | **A4/LEARN**<br>**N = 171<sup>1</sup>** | **ADNI**<br>**N = 114<sup>1</sup>** | **HABS**<br>**N = 31<sup>1</sup>** |
| --- | --- | --- | --- | --- |
| Age | 73 (7)<br>[57, 94] | 72 (5)<br>[65, 86] | 74 (8)<br>[57, 94] | 78 (7)<br>[66, 92] |
| Years of Education | 15.81 (2.62)<br>[9.00, 22.00] | 15.56 (2.75)<br>[9.00, 22.00] | 16.20 (2.35)<br>[12.00, 20.00] | 15.81 (2.77)<br>[11.00, 20.00] |
| Race/ ethnicity |  |  |  |  |
| Hispanic or Latino | 15 (4.7%) | 5 (2.9%) | 9 (7.9%) | 1 (3.2%) |
| Not Hispanic or Latino | 296 (94%) | 162 (95%) | 104 (91%) | 30 (97%) |
| Unknown | 5 (1.6%) | 4 (2.3%) | 1 (0.9%) | 0 (0%) |
| APOEε4 (ε4+) | 167 (55%) | 95 (56%) | 51 (50%) | 21 (70%) |
| Unknown | 14 | 0 | 13 | 1 |
| Baseline MTL tau | 1.18 (0.13)<br>[0.87, 1.62] | 1.17 (0.12)<br>[0.87, 1.58] | 1.19 (0.14)<br>[0.90, 1.62] | 1.24 (0.13)<br>[1.06, 1.52] |
| Baseline neocortical tau | 1.17 (0.08)<br>[0.93, 1.35] | 1.16 (0.08)<br>[0.93, 1.35] | 1.20 (0.07)<br>[1.01, 1.35] | 1.19 (0.07)<br>[1.06, 1.35] |
| Baseline Amyloid | 56 (28)<br>[6, 160] | 56 (28)<br>[6, 160] | 53 (28)<br>[18, 137] | 62 (29)<br>[24, 126] |
| Unknown | 6 | 3 | 2 | 1 |
| MMSE | 28.82 (1.53)<br>[14.00, 30.00] | 28.88 (1.17)<br>[24.00, 30.00] | 28.62 (2.07)<br>[14.00, 30.00] | 29.16 (1.07)<br>[27.00, 30.00] |
| Unknown | 12 | 0 | 12 | 0 |
| IPACC | 0.64 (0.48)<br>[-1.72, 2.00] | 0.71 (0.37)<br>[-0.07, 2.00] | 0.46 (0.57)<br>[-1.72, 1.72] | 0.89 (0.49)<br>[-0.12, 1.90] |
| Unknown | 13 | 0 | 12 | 1 |
| Resistance_corrected | -0.23 (0.31)<br>[-1.31, 0.58] | -0.22 (0.30)<br>[-1.31, 0.57] | -0.26 (0.35)<br>[-1.28, 0.58] | -0.17 (0.25)<br>[-0.61, 0.26] |

<sup>1</sup>Mean (SD) [Min, Max]; n (%)

**Figure 1.**
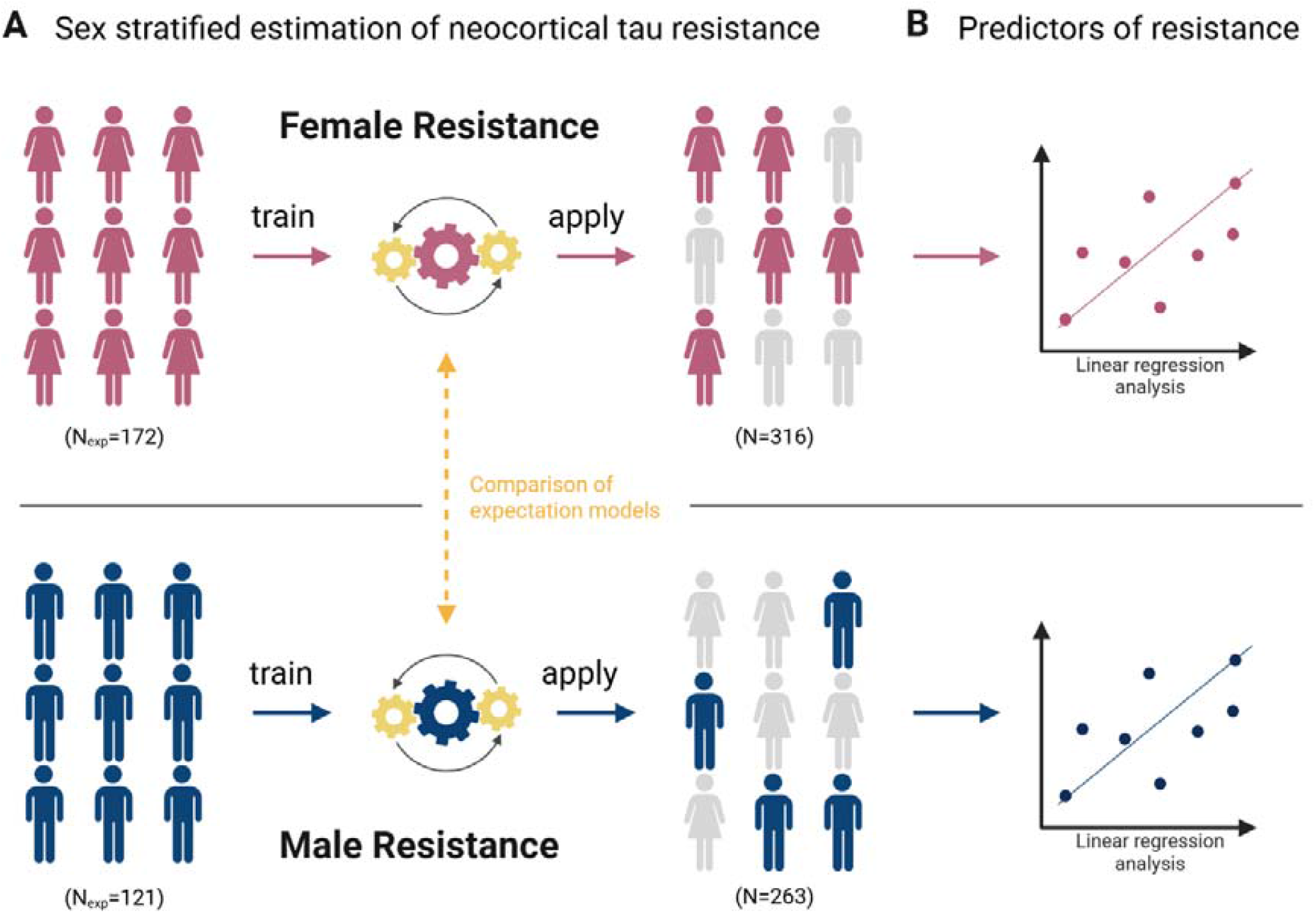
Study Overview. Operationalization of sex-specific resistance to neocortical tau. Training two separate expectation models in male (N_train_=121) and female populations (N_Train_=172) that experienced a more typical clinical progression during their lives and subsequent application of each trained model on a heterogeneous population (N_Test_=579) stratified by men (N=263) and women (N=316) to estimate resistance (A), and comparison of expectation models in terms of model feature importance (highlighted in yellow). Investigating predictors male and female resistance to neocortical tau within each model using linear regression analyses (B). Created with BioRender.com.

### 2.6 Statistical Analyses

Statistical analysis was performed in R version 4.3.2 (The R Foundation, Vienna, Austria). To assess differences in female-only and male-only expectation models, we first compared the feature importance of the same predictors in the female-only and male-only expectation models. For this, we calculated the 95% confidence intervals for the difference in means in feature importance between both models per variable across 1000 bootstrap samples. As such, feature importance was quantified using the variables coefficients in the model. We then investigated potential predictors (age, cognition, education, MTL tau, ICV-adjusted hippocampal volume, and amyloid burden) of tau resistance within each sex. For this, we ran a series of linear regression models in women using resistance scores estimated by our female-only expectation model, and in a separate series, in men using resistance scores estimated by our male-only expectation model. In each of these analyses, predictors were standardized before fitting the model and covariates were age, education, *APOE*ε4 status, and cohort.

## 3. RESULTS

### 3.1 Sex-specific model feature importance

When assessing the concordance of the most important (top 5 positive and top 5 negative, **Figures 2-A** and **2-B**) features in the female-only and male-only models, we found that only 4 of them were overlapping (see also **Supplementary Figures 1** and **2** for all features). Cognition, as measured using the global CDR score and lPACC, as well as age and race/ethnicity, were represented as important model features in both models (**Figures 2-C** and **Supplementary Figure 3**). Interestingly, the race/ethnicity variable impacted the sex-specific models in opposite directions. Among those top 10, features distinctively important to the male-only model included Mini Mental State Examination (MMSE) and Aβ, whereas *APOE*ε4 contributed considerably to the female-only model. Comparing the mean bootstrapped feature importance across the female-only and male-only model revealed statistically significant differences for 148 out of the total 153 variables (140 of which are MRI-related; **Figure 2-D** and **Supplementary Figure 4**, See **Supplementary Table 2**). Specifically, race/ethnicity (Importance in male-only model, I_m_=0.03, Importance in female-only model, I_f_=-0.17, Difference in means, D_*M*_=0.20, CI[0.19,0.21]), *APOE*ε4 (I_m_=0.02, I_f_=-0.08, D_*M*_=0.10, CI[0.05,0.05]), lPACC (I_m_=-0.08, I_f_=-0.15, D_*M*_=0.07, CI[0.07,0.08]), smoking (I_m_=0.01, I_f_=-0.03, D_*M*_=0.03, CI[0.03,0.04]), education (I_m_=-0.00, I_f_=0.02, D_*M*_=0.02, CI[0.02,0.02]), and global CDR (I_m_=0.05, I_f_=0.05, D_*M*_=0.00, CI[0.00,0.01]) weighted higher in the female-only as compared to male-only expectation model. By contrast, MMSE (I_m_=-0.08, I_f_=0.01, D_*M*_=0.09, CI[0.09,0.09]), age, (I_m_=-0.14, I_f_=-0.09, D_*M*_=0.05, CI[0.05,0.05]), Aβ?(I_m_=0.05, I_f_=-0.00, D_*M*_=0.05, CI[0.05,0.05]), as well as metabolic measures including diastolic and systolic blood pressure, (I_m_=0.01, I_f_=-0.00, D_*M*_=0.02, CI[0.02,0.02] and I_m_=-0.01, I_f_=0.00, D_*M*_=0.02, CI[0.02,0.01], respectively), body weight (I_m_=-0.01, I_f_=0.00, D_*M*_=0.01, CI[0.01,0.01]), and pulse (I_m_=0.01, I_f_=-0.00, D_*M*_=0.01, CI[0.01,0.01]) weighted significantly higher in the male-only expectation model as compared to the female-only expectation model.

**Table 2.**
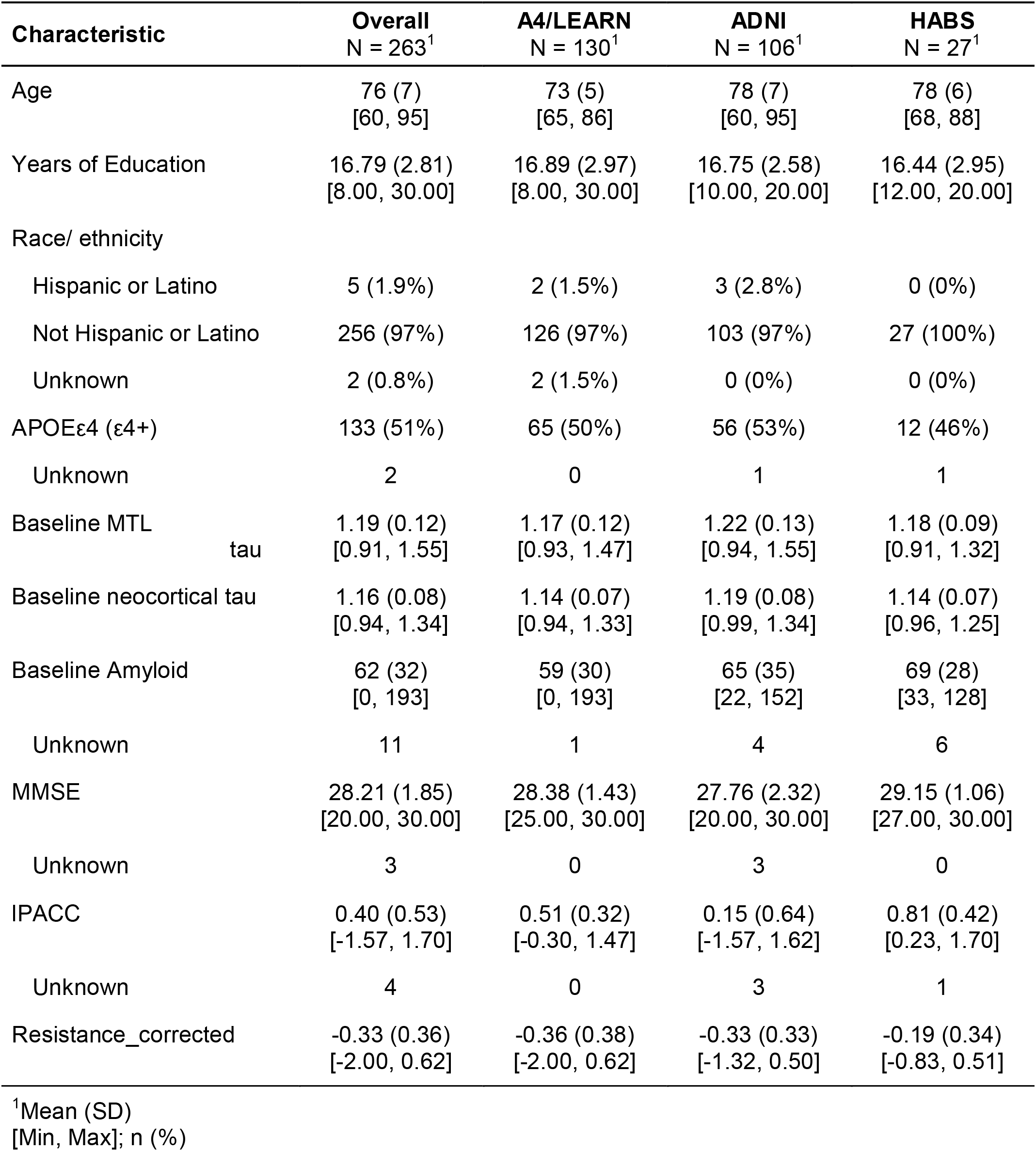
Male Sample Demographics.

**Figure 2.**
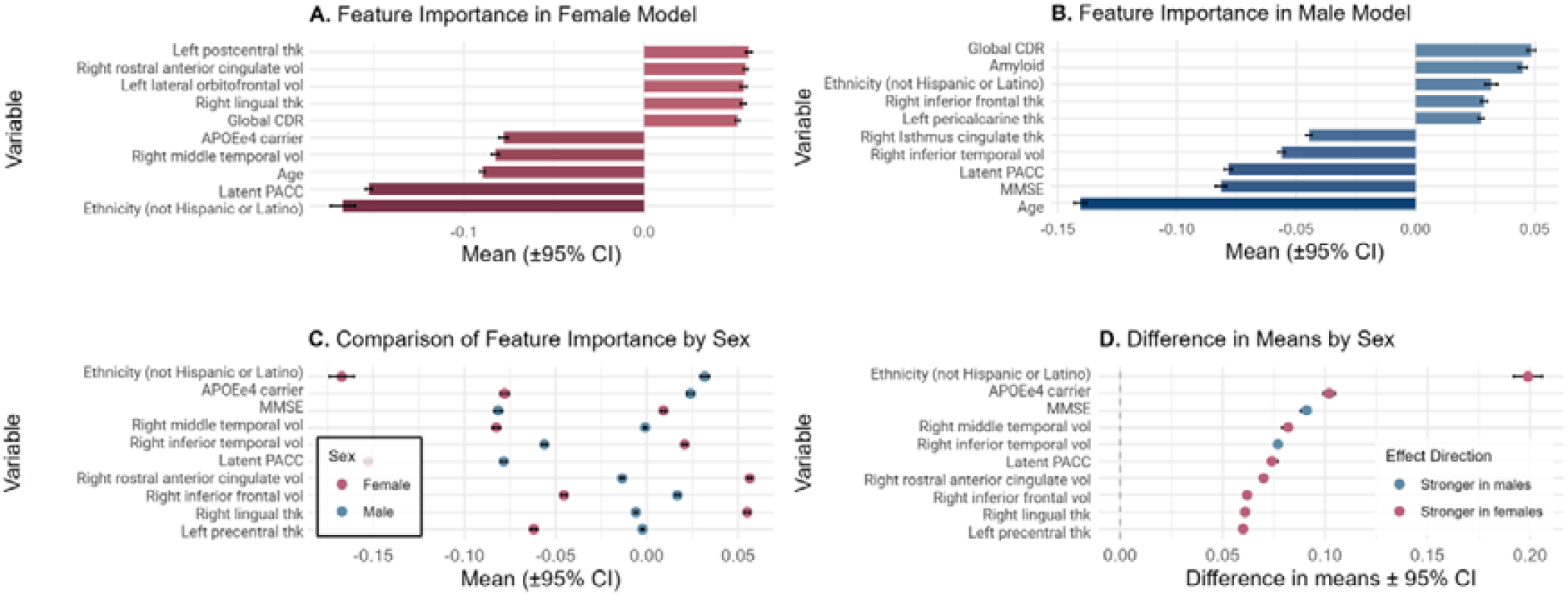
Sex Differences in Feature Importance for Sex-specific Expectation Models. Top 10 mean bootstrapped estimates of feature importance (±95% confidence interval, Top 5 positive and Top 5 negative) in sex-stratified expectation models are shown separately for females (A) and males (B), ordered by descending mean importance of features. Direct comparison of top 10 mean bootstrapped feature importance (±95% confidence interval) from male and female models plotted jointly for each feature for visual comparison of sex-specific model contributions (C). Difference in means between female and male feature weights of top 10 (highest magnitude) variables (±95% confidence interval) are shown in (D), reflecting the relative strength and direction of sex effects on model feature importance. Values in blue indicate greater importance in the male model, while pink values indicate greater importance in the female model.

**Figure 3.**
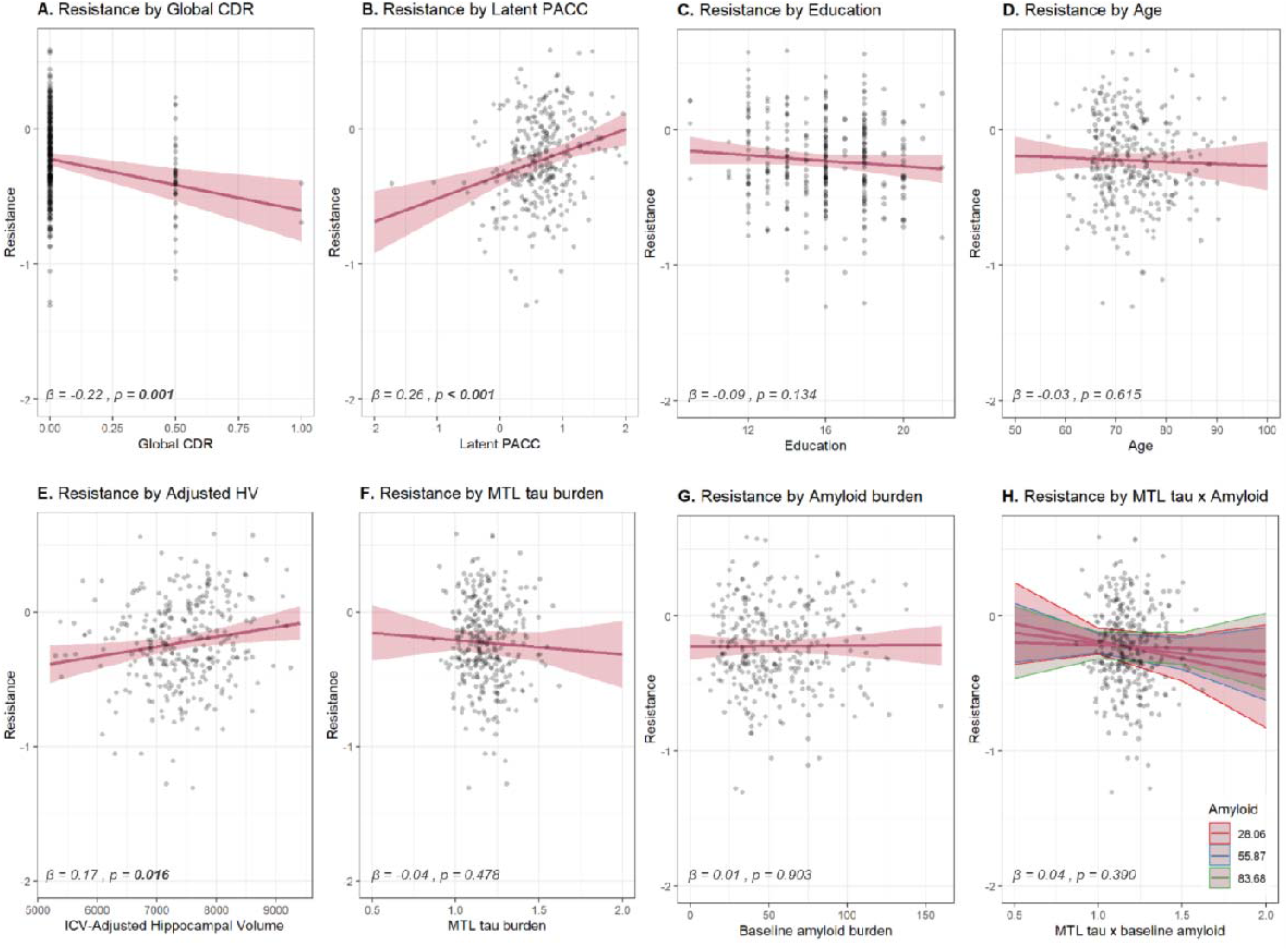
Predictors of Female Resistance to Neocortical Tau. Higher global CDR associated with lower necortical tau resistance in women (A. β=-0.22, p<0.01), whereas higher latent PACC was predictive of higher female resistance (B. β=0.26, p<0.001), as was higher ICV-adjusted hippocampal volume (E. β=0.17, p<0.05). Female resistance was not predicted by years of education (C. β=-0.09, p=0.134), age (D. β=-0.03, p=0.615), MTL tau (F. β=-0.04, p=0.476) or amyloid burden (G. β=0.01, p=0.903), nor by the interaction of MTL tau and amyloid burden (H. β=0.04, p=0.390). Effect sizes are reported as standardized betas (β) to allow comparison across predictors.

**Figure 4.**
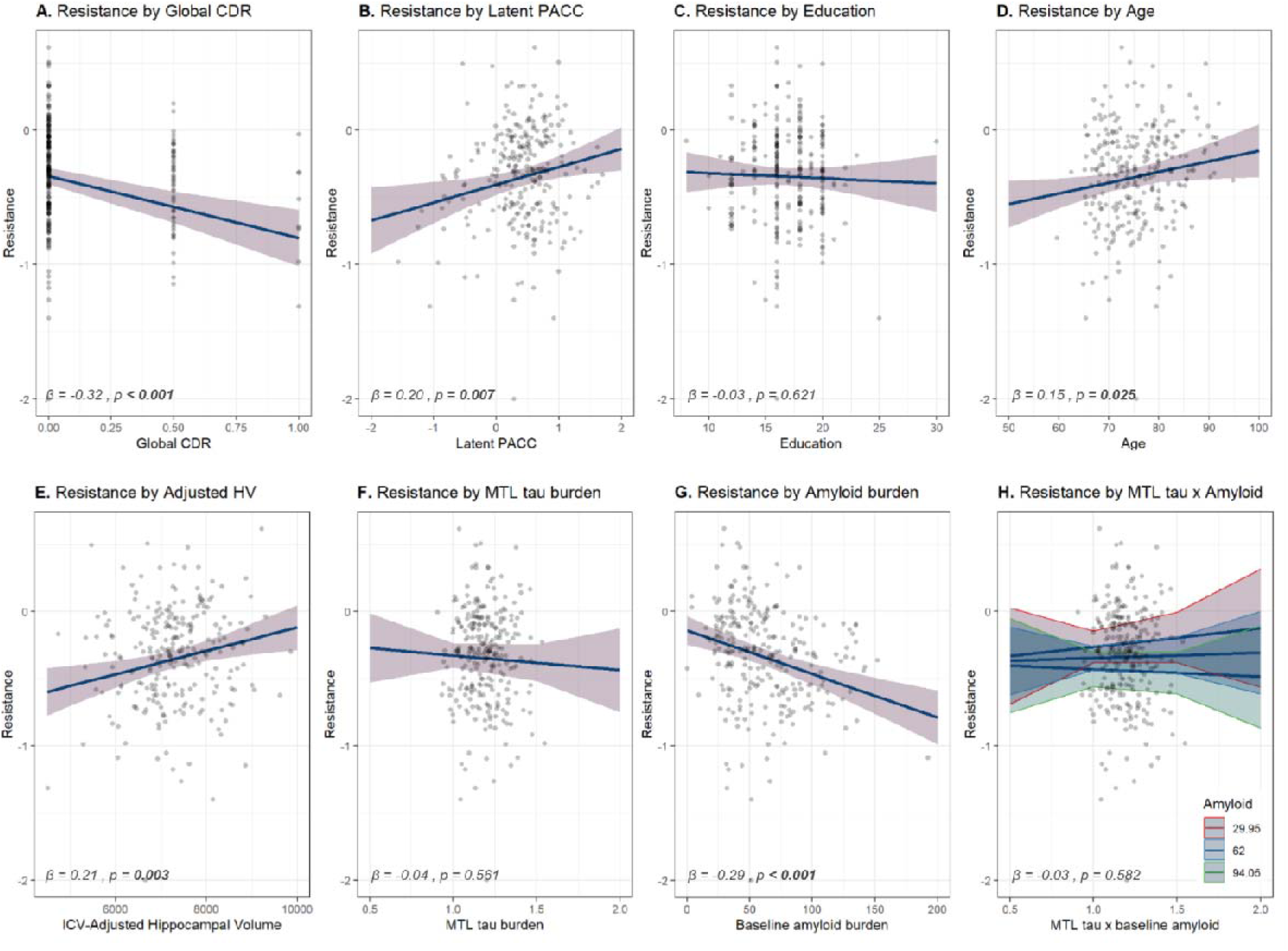
Predictors of Male Resistance to Neocortical Tau. Higher global CDR (A. β=-0.32, p<0.001) and baseline amyloid burden (G. β=-0.29, p<0.001) were predictive of lower male necortical tau resistance, whereas latent PACC (B. β=0.20, p<0.01), age (D. β=0.15, p<0.05) and ICV-adjusted hippocampal volume (E. β=0.21, p<0.05) were predictive of higher male resistance. In Men, resistance was not predicted by years of education (C. β=-0.03, p=0.621), MTL tau (F. β=-0.04, p=0.561), or the interaction of MTL tau and amyloid burden (H. β=-0.03, p=0.582). Effect sizes are reported as standardized betas (β) to allow comparison across predictors.

### 3.2 Female resistance

Higher global CDR scores, reflecting worse cognition, were associated with lower female resistance (**Figure 3-A**; β=-0.22, CI[-0.35,-0.09], p<0.01) while higher latent PACC, reflecting better cognition, associated with higher female resistance (**Figure 3-B**; β=0.26, CI[0.13,0.39], p<0.001). Hippocampal volume was significantly associated with higher female resistance (**Figure 3-E**; β=0.17, CI[0.03,0.30], p<0.05). By contrast, education, age, MTL tau and Aβ-PET burden were not associated with female NEO tau resistance (see **Figures 3-C, D, F, G**, respectively). Finally, we found no interaction between a MTL tau and Aβ on female NEO tau resistance (**Figure 3-H**).

### 3.3 Male resistance

Higher global CDR scores associated with lower male resistance (**Figure 4-A**; β=-0.32, CI[-0.47,-0.18], p<0.001), and higher latent PACC were associated with higher male resistance (**Figure 4-B**; β=0.20, CI[0.05,0.34], p<0.01). Higher age was associated with higher male resistance (**Figure 4-D**; β=0.15, CI[0.02,0.28], p<0.05), as was ICV-adjusted hippocampal volume (**Figure 4-E**; β=0.21, CI[0.07,0.35], p<0.01). By contrast, higher Aβ-PET burden was associated with lower male tau resistance (Figure 4-G; β=-0.29, CI[-0.41,-0.16], p<0.001). We found no association between male resistance and education (**Figure 4-C**) or MTL tau (**Figure 4-F**). We found no significant association of an MTL tau and amyloid interaction with male resistance (**Figure 4-H**).

## 4. DISCUSSION

Sex differences to AD pathology have been established extensively across various disease-related factors including Aβ-associated tau burden [5], cognitive decline [28], and AD progression [29]. In this study we employed our recently published inverse learning approach to estimate NEO tau resistance in a sample of female and male individuals who represent caTAUstrophe, defined as the detrimental spread of tau from MTL to NEO regions [4]. Specifically, we built two separate sex-specific models predicting NEO tau burden and applied each model to a sample of women and men, respectively, who potentially included individuals with higher levels of tau resistance. We found that almost all variables differed in their importance for predicting NEO tau resistance between the sex-specific models. Most notably, cognition (both latent PACC and global CDR), education, and *APOE*ε4 weighted stronger in the female-only model, while age and Aβ weighted more strongly in the male-only model. Our findings align with well-established sex differences in AD, particularly with regard to sex differences in susceptibility to tau accumulation and disease progression [28,29], highlighting the importance of sex-related characteristics in each sex when predicting disease progression. However, it is important to note that in this study, resistance overall was quite low in both men and women, with most individuals’ resistance estimates placing below zero. Interestingly, male resistance in particular presented with a higher skew towards lower resistance. These findings, in combination with previous evidence on sex-differences to AD-related pathophysiology, further highlight the need for sex-specific operationalization of NEO tau resistance.

When investigating sex-specific and sex-agnostic predictors of resistance, we found that higher global CDR and lower latent PACC scores were predictive of lower NEO tau resistance in both sexes, indicating that individuals with cognitive impairment were less resistant to caTAUstrophe. Interestingly, years of education – often defined as a proxy measure of cognitive resilience (or better than expected cognitive performance in the context of AD [30]) – did not predict resistance in either sex. Together, these findings suggest that 1) cognition is predictive of NEO tau resistance in both women and men, and 2) educational attainment does not predict NEO tau resistance in either sex in our sample. This is in line with a study by Ramanan and colleagues [31], who found no association between education and regional tau burden. Together, these findings importantly distinguish resistance (avoiding pathology) conceptually from cognitive and brain resilience (coping with pathology), for which education is the most robust determinant[30,32,33].

While hippocampal volume predicted both female and male NEO tau resistance, indicating a greater involvement of the hippocampus in brain reserve specific to NEO tau resistance, older age was associated with higher male but not female resistance, suggesting that age is uniquely predictive of NEO tau resistance in men. Moreover, concordant with previous findings highlighting different rates of Aβ-induced acceleration of caTAUstrophy in women and men, and a female disadvantage in the brain’s ability to cope with Aβ-proteinopathy[5], Aβ burden at baseline was predictive of male resistance but not female resistance. MTL tau burden did not predict resistance in either sex, suggesting that baseline MTL tau might not be a sufficient indicator of NEO tau resistance at least cross-sectionally, despite posing a higher risk for caTAUstrophe[4,34]. In addition, the differential relationship between amyloid burden and sex-specific resistance was not modulated by MTL tau in either sex, supporting this female disadvantage downstream of initial amyloid and tau deposition.

Taken together, our study highlights existing sex-differences in the predictors of NEO tau resistance. Nonetheless, we acknowledge several limitations. First, estimating NEO tau resistance using our inverse learning approach is influenced by the characteristics of the expectation sample employed for training the predictive model. Running these analysis with a different expectation sample could yield differing results. However, we aimed to have a robust expectation sample by pooling several data-rich cohorts. Furthermore, sex-stratification of both the expectation sample and our test sample resulted in relatively small sample sizes, hence reducing statistical power that potentially limited the identification of relevant associations. Finally, while our study was cross-sectional, we aim to conduct longitudinal studies of sex differences in NEO tau resistance in future work given its dynamic nature.

In conclusion, we demonstrate that resistance to AD-related NEO tau burden is systematically sex-driven in both its expectation and prediction. Therefore, we strongly advise the use of sex-specific models, as crucial sex-specific characteristics are likely to “get cancelled out” when using pooled-sex approaches. A better understanding of sex-specific pathways that promote resistance to NEO tauopathy is necessary to identify protective factors that may be leveraged for targeted interventions.

## Data Availability

De-identified individual-level data and aggregate-level statistics from ADNI, HABS, and A4/LEARN are available to registered researchers, with access provided through the respective study portals (ADNI via the LONI IDA, HABS via the HABS data portal, and A4/LEARN via Synapse) under data use agreements.

## Abbreviations

Aβ: amyloid-beta
T: Tau
NEO: Neocortical
AD: Alzheimer’s disease
APOE: Apolipoprotein
MRI: magnetic resonance imaging
PET: positron emission tomography
ICV: intracranial volume
SUVr: standardized uptake volume ratio
FBB: Florbetaben
FBP: 18F- Florbetapir
PiB: 11C Pittsburgh compound-B
ROI: region of interest
(l)PACC: (Latent) Preclinical Alzheimer’s Cognitive Composite
CDR: Clinical Dementia Rating
DVR: distribution volume ratio
A4/LEARN: Longitudinal Evaluation of Amyloid Risk and Neurodegeneration study
HABS: Harvard Aging Brain Study
ADNI: Alzheimer’s Disease Neuroimaging Initiative

## ACKNOWLEDGEMENTS

We thank the participants of the studies for their contribution to this important research.

## CONSENT STATEMENT

This study complies with the declaration of Helsinki. All participants provided written informed consent.

## Notes

### Competing Interest Statement

T.J.H. is on the Scientific Advisory Board for Circular Genomics, serves as Deputy Editor for Alzheimer's & Dementia: Translational Research and Clinical Intervention, and serves as Section Editor for Alzheimer's & Dementia. R.O. is currently a full-time employee of Eli Lilly and Company. His contribution to the work presented in this manuscript was performed as an employee of Amsterdam University Medical Centers. R.O. has received research funding/support from European Research Council, ZonMw, NWO, National Institute of Health, Alzheimer Association, Alzheimer Nederland, Stichting Dioraphte, Cure Alzheimer's fund, Health Holland, ERA PerMed, Alzheimerfonden, Hjarnfonden, Avid Radiopharmaceuticals, Janssen Research & Development, Roche, Quanterix and Optina Diagnostics, has given lectures in symposia sponsored by GE Healthcare, received speaker fees from Springer, and was an advisory board/steering committee member for Asceneuron, Biogen, Johnson & Johnson and Bristol Myers Squibb. All the aforementioned has been paid to Amsterdam University Medical Centers. All other authors have nothing to disclose.

### Funding Statement

This research is funded by Alzheimer Nederland (WE.08-2024-06 and WE.03-2021-03).
M.S. is supported by a research fellowship from the Alzheimer's Association (24AARF-1201281). G.T.C. is funded on the National Institute on Aging (K99 AG083063), as well as a research fellowship from the Alzheimer's Association (AARF-23-1151259). T.J.H. has received funding from U24AG074855, R01AG079142, R01AG073439, and U01AG082350. R.O. has received funding from the European Research Council (ERC) under the European Union's Horizon 2020 research and innovation programme (grant agreement No. 949570). R.F.B. is supported by the National Institute on Aging (R01AG079142 and DP2 AG082342).

### Author Declarations

De-identified individual-level data and aggregate-level statistics from ADNI, HABS, and A4/LEARN are available to registered researchers, with access provided through the respective study portals (ADNI via the LONI IDA, HABS via the HABS data portal, and A4/LEARN via Synapse) under data use agreements. We conducted this study under the ethical guidelines stipulated by the Partners Human Research Committee, which is the Institutional Review Board for the Massachusetts General Hospital and Brigham and Women's Hospital.

