## Supplementary for "Predictors of sex-specific resistance to caTAUstrophe"

**SUPPLEMENTARY MATERIAL**

**Definition of Tau Positivity**

To determine a cut-off for elevated, abnormal levels of tau pathology in the neocortical brain regions (i.e., neocortical tau positivity), we used Gaussian Mixture Models (GMM). We fitted a GMM with two components to our calculated neocortical composite scores of our full dataset. The cut-off was then placed between the two components of the GMM. Furthermore, we compared the fitted two-component GMM against a fitting a single normal distribution using the Bayesian information criteria (BIC), to ensure that the data supports a two-component model. The code can be found at: https://github.com/Cojabi/gmmthresholder.

**Supplementary Figure 1 Feature Importance in Female Model**

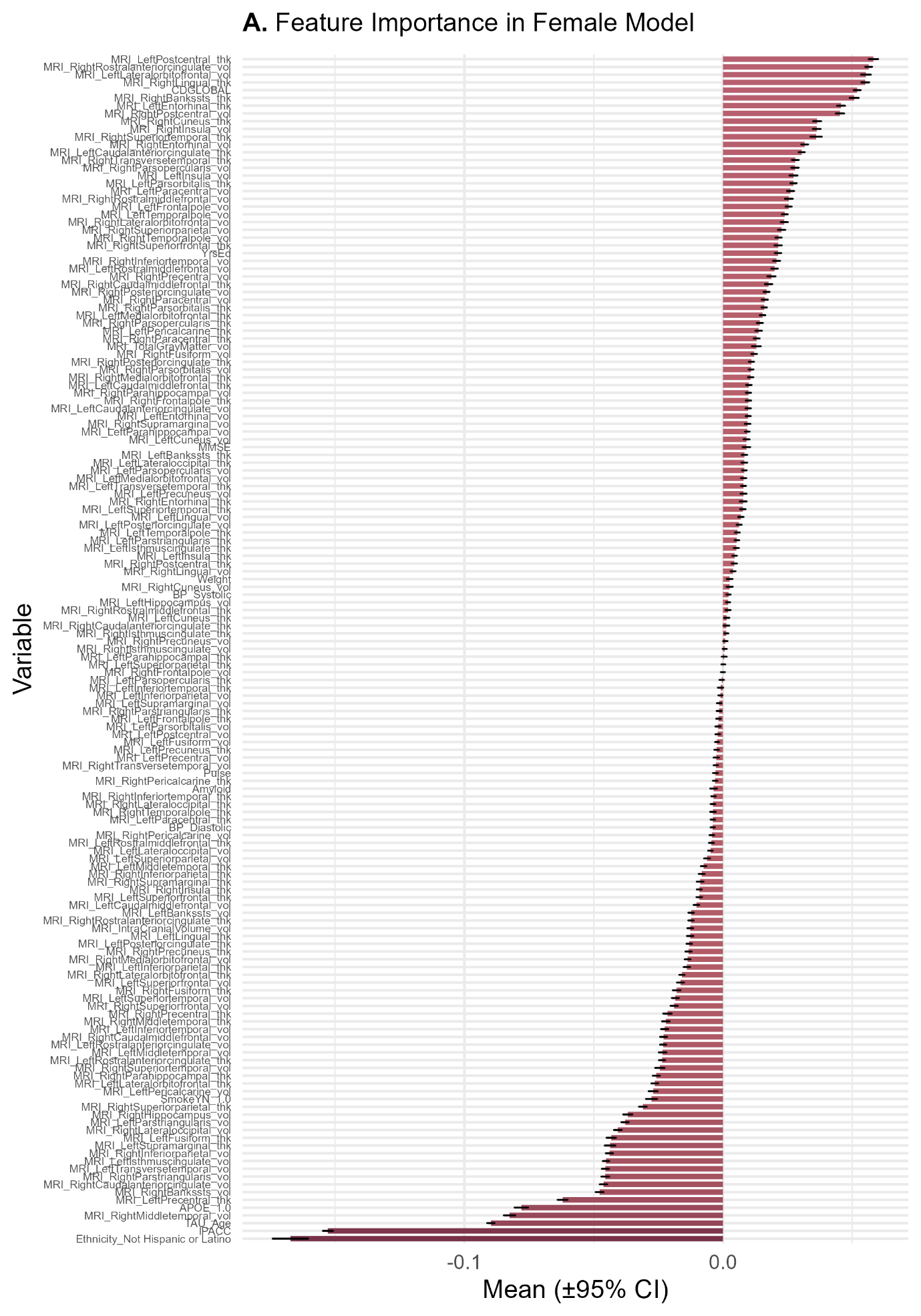

**Supplementary Figure 2 Feature Importance in Male Model**

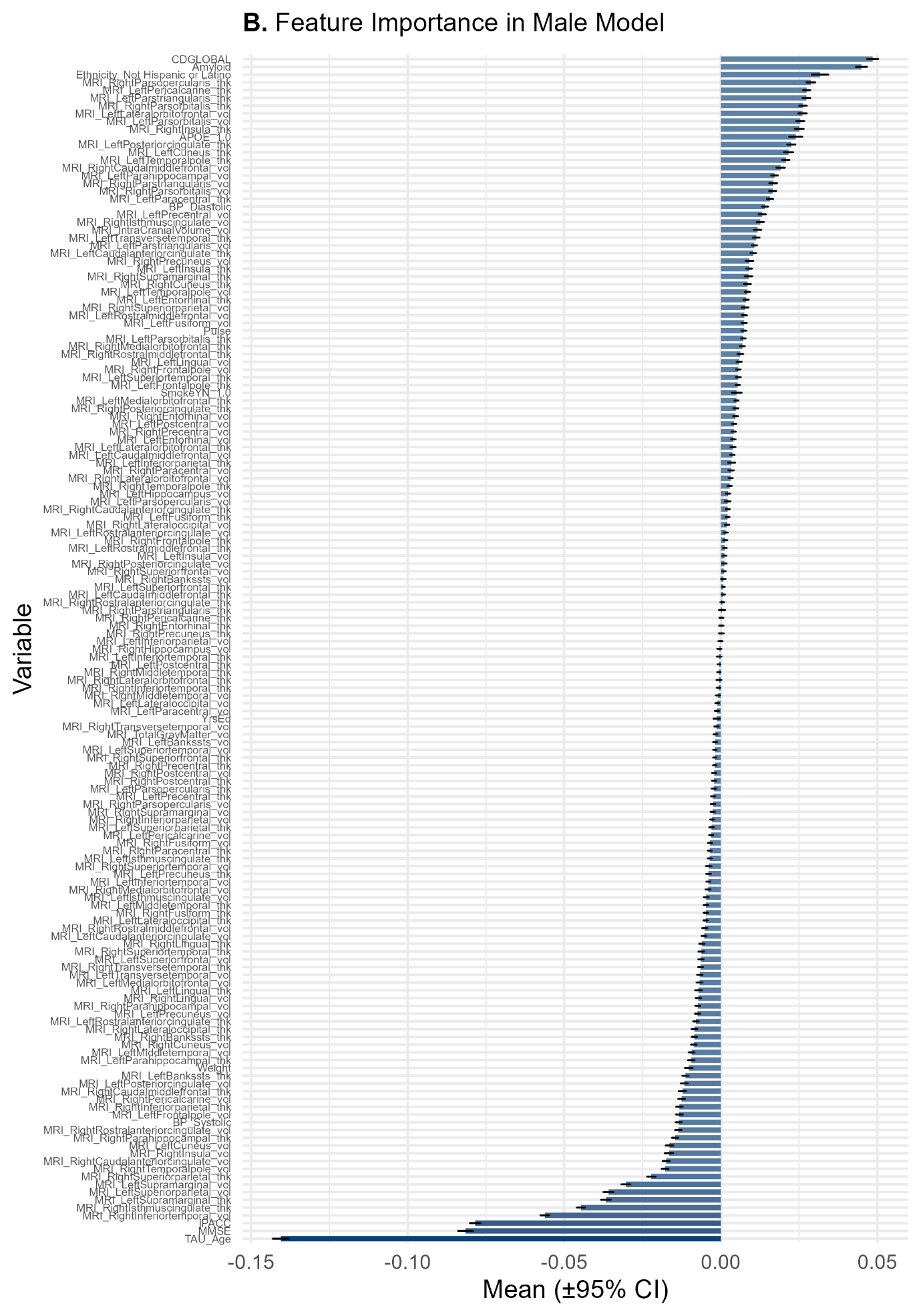

**Supplementary Figure 3 Comparison of Feature Importance by Sex**

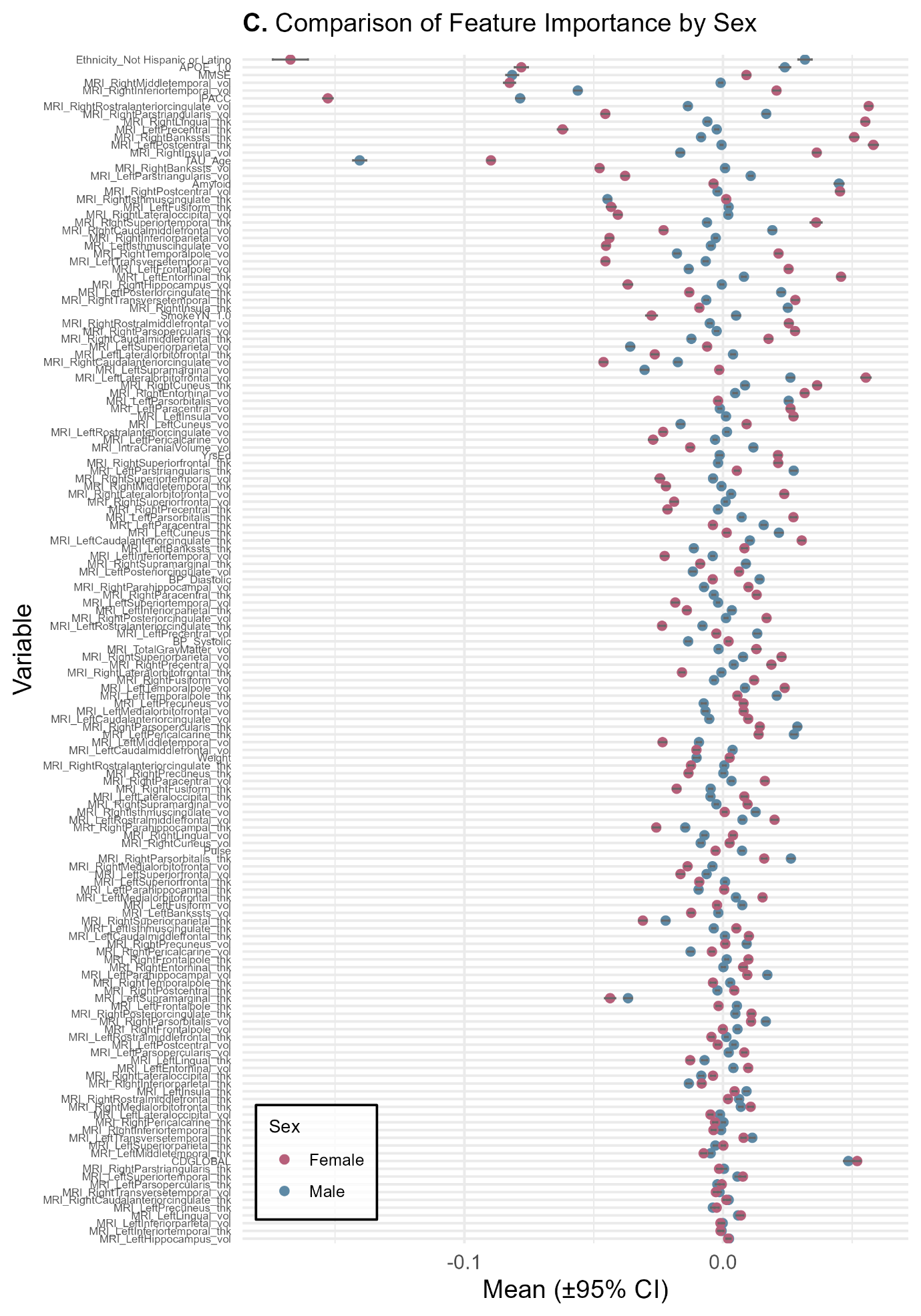

**Supplementary Figure 4 Difference in Means by Sex**

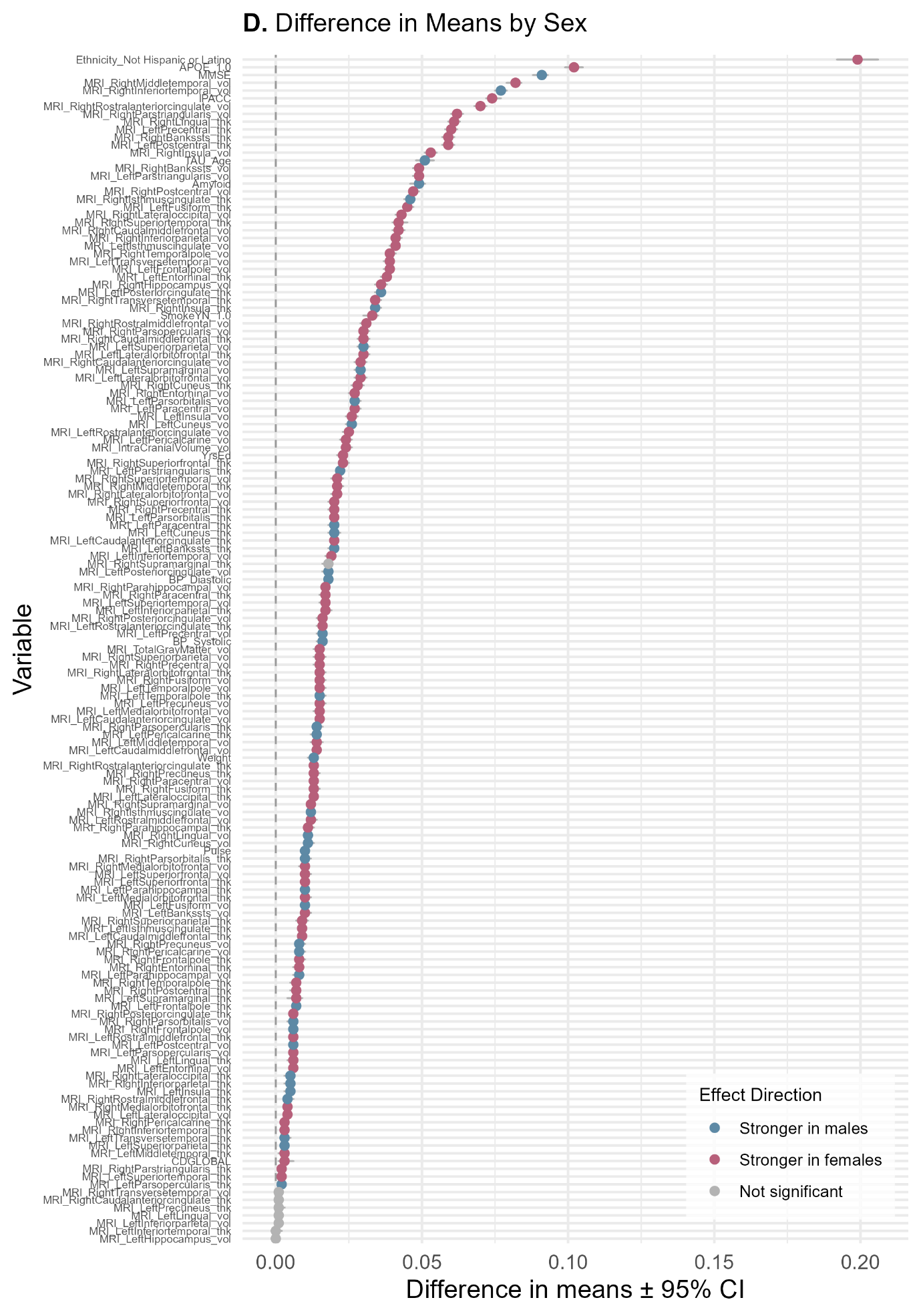

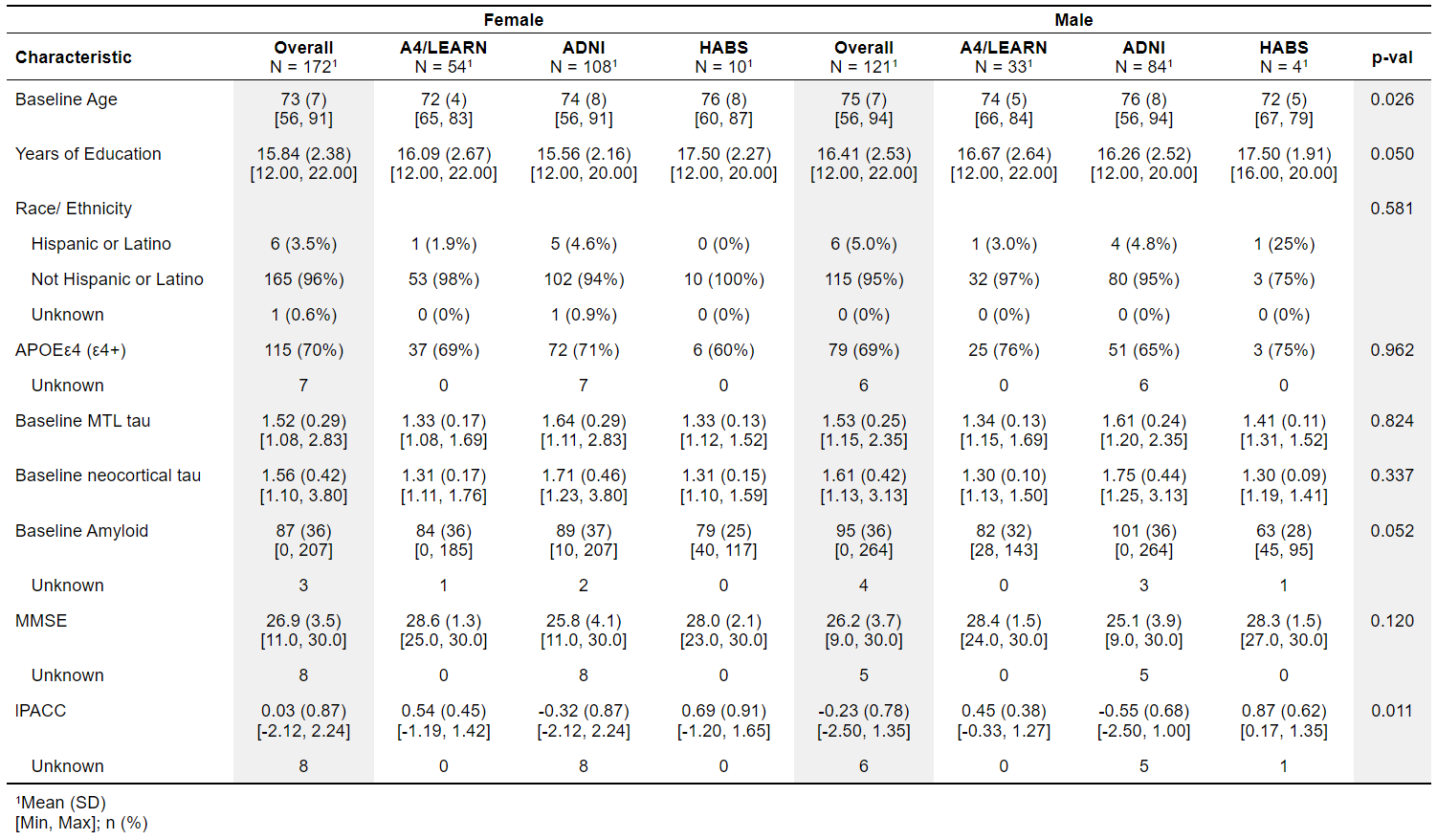
**Supplementary Table 1 Expectation samples**

**Supplementary Table 2 Difference in mean feature weights between Models**

| **Variable** | **Mean (Male)** | **Mean (Female)** | **Difference in Means** | **Lower 95% CI** | **Upper 95% CI** | **Significant (95% CI excludes 0)** |
| --- | --- | --- | --- | --- | --- | --- |
| APOE_1.0 | 0.024 | -0.078 | 0.102 | 0.099 | 0.105 | TRUE |
| Amyloid | 0.045 | -0.004 | 0.049 | 0.046 | 0.051 | TRUE |
| BP_Diastolic | 0.014 | -0.004 | 0.018 | 0.017 | 0.019 | TRUE |
| BP_Systolic | -0.013 | 0.002 | 0.016 | 0.014 | 0.017 | TRUE |
| CDGLOBAL | 0.048 | 0.052 | 0.003 | 0.001 | 0.006 | TRUE |
| Ethnicity_Not Hispanic or Latino | 0.032 | -0.167 | 0.199 | 0.192 | 0.206 | TRUE |
| MMSE | -0.082 | 0.009 | 0.091 | 0.088 | 0.093 | TRUE |
| MRI_IntraCranialVolume_vol | 0.012 | -0.013 | 0.024 | 0.023 | 0.026 | TRUE |
| MRI_LeftBankssts_thk | -0.011 | 0.008 | 0.020 | 0.018 | 0.021 | TRUE |
| MRI_LeftBankssts_vol | -0.002 | -0.012 | 0.010 | 0.009 | 0.012 | TRUE |
| MRI_LeftCaudalanteriorcingulate_thk | 0.010 | 0.030 | 0.020 | 0.019 | 0.021 | TRUE |
| MRI_LeftCaudalanteriorcingulate_vol | -0.005 | 0.010 | 0.015 | 0.014 | 0.016 | TRUE |
| MRI_LeftCaudalmiddlefrontal_thk | 0.001 | 0.010 | 0.009 | 0.008 | 0.010 | TRUE |
| MRI_LeftCaudalmiddlefrontal_vol | 0.004 | -0.010 | 0.014 | 0.013 | 0.015 | TRUE |
| MRI_LeftCuneus_thk | 0.022 | 0.001 | 0.020 | 0.018 | 0.022 | TRUE |
| MRI_LeftCuneus_vol | -0.016 | 0.009 | 0.026 | 0.024 | 0.027 | TRUE |
| MRI_LeftEntorhinal_thk | 0.008 | 0.046 | 0.038 | 0.036 | 0.039 | TRUE |
| MRI_LeftEntorhinal_vol | 0.004 | 0.010 | 0.006 | 0.005 | 0.007 | TRUE |
| MRI_LeftFrontalpole_thk | 0.005 | -0.002 | 0.007 | 0.006 | 0.008 | TRUE |
| MRI_LeftFrontalpole_vol | -0.013 | 0.025 | 0.039 | 0.037 | 0.040 | TRUE |
| MRI_LeftFusiform_thk | 0.002 | -0.043 | 0.045 | 0.044 | 0.047 | TRUE |
| MRI_LeftFusiform_vol | 0.007 | -0.002 | 0.010 | 0.009 | 0.011 | TRUE |
| MRI_LeftHippocampus_vol | 0.002 | 0.002 | 0.000 | -0.001 | 0.001 | FALSE |
| MRI_LeftInferiorparietal_thk | 0.003 | -0.014 | 0.017 | 0.016 | 0.019 | TRUE |
| MRI_LeftInferiorparietal_vol | 0.000 | -0.001 | 0.001 | 0.000 | 0.002 | FALSE |
| MRI_LeftInferiortemporal_thk | -0.001 | -0.001 | 0.000 | -0.001 | 0.002 | FALSE |
| MRI_LeftInferiortemporal_vol | -0.004 | -0.023 | 0.019 | 0.017 | 0.020 | TRUE |
| MRI_LeftInsula_thk | 0.009 | 0.004 | 0.005 | 0.003 | 0.006 | TRUE |
| MRI_LeftInsula_vol | 0.001 | 0.027 | 0.026 | 0.024 | 0.028 | TRUE |
| MRI_LeftIsthmuscingulate_thk | -0.004 | 0.005 | 0.009 | 0.008 | 0.010 | TRUE |
| MRI_LeftIsthmuscingulate_vol | -0.005 | -0.045 | 0.041 | 0.039 | 0.042 | TRUE |
| MRI_LeftLateraloccipital_thk | -0.005 | 0.008 | 0.013 | 0.012 | 0.014 | TRUE |
| MRI_LeftLateraloccipital_vol | -0.001 | -0.005 | 0.004 | 0.003 | 0.005 | TRUE |
| MRI_LeftLateralorbitofrontal_thk | 0.004 | -0.026 | 0.030 | 0.029 | 0.032 | TRUE |
| MRI_LeftLateralorbitofrontal_vol | 0.026 | 0.055 | 0.029 | 0.027 | 0.031 | TRUE |
| MRI_LeftLingual_thk | -0.007 | -0.013 | 0.006 | 0.004 | 0.007 | TRUE |
| MRI_LeftLingual_vol | 0.006 | 0.007 | 0.001 | 0.000 | 0.002 | FALSE |
| MRI_LeftMedialorbitofrontal_thk | 0.005 | 0.015 | 0.010 | 0.009 | 0.012 | TRUE |
| MRI_LeftMedialorbitofrontal_vol | -0.007 | 0.008 | 0.015 | 0.013 | 0.016 | TRUE |
| MRI_LeftMiddletemporal_thk | -0.005 | -0.007 | 0.003 | 0.001 | 0.004 | TRUE |
| MRI_LeftMiddletemporal_vol | -0.009 | -0.023 | 0.014 | 0.012 | 0.016 | TRUE |
| MRI_LeftParacentral_thk | 0.016 | -0.004 | 0.020 | 0.018 | 0.021 | TRUE |
| MRI_LeftParacentral_vol | -0.001 | 0.026 | 0.027 | 0.026 | 0.029 | TRUE |
| MRI_LeftParahippocampal_thk | -0.009 | 0.000 | 0.010 | 0.009 | 0.011 | TRUE |
| MRI_LeftParahippocampal_vol | 0.017 | 0.009 | 0.008 | 0.006 | 0.009 | TRUE |
| MRI_LeftParsopercularis_thk | -0.002 | 0.000 | 0.002 | 0.001 | 0.003 | TRUE |
| MRI_LeftParsopercularis_vol | 0.002 | 0.008 | 0.006 | 0.005 | 0.007 | TRUE |
| MRI_LeftParsorbitalis_thk | 0.007 | 0.027 | 0.020 | 0.019 | 0.021 | TRUE |
| MRI_LeftParsorbitalis_vol | 0.025 | -0.002 | 0.027 | 0.026 | 0.029 | TRUE |
| MRI_LeftParstriangularis_thk | 0.027 | 0.005 | 0.022 | 0.021 | 0.023 | TRUE |
| MRI_LeftParstriangularis_vol | 0.011 | -0.038 | 0.049 | 0.047 | 0.050 | TRUE |
| MRI_LeftPericalcarine_thk | 0.027 | 0.014 | 0.014 | 0.012 | 0.015 | TRUE |
| MRI_LeftPericalcarine_vol | -0.003 | -0.027 | 0.024 | 0.022 | 0.026 | TRUE |
| MRI_LeftPostcentral_thk | -0.001 | 0.058 | 0.059 | 0.057 | 0.061 | TRUE |
| MRI_LeftPostcentral_vol | 0.004 | -0.002 | 0.006 | 0.005 | 0.007 | TRUE |
| MRI_LeftPosteriorcingulate_thk | 0.023 | -0.013 | 0.036 | 0.034 | 0.037 | TRUE |
| MRI_LeftPosteriorcingulate_vol | -0.012 | 0.006 | 0.018 | 0.016 | 0.019 | TRUE |
| MRI_LeftPrecentral_thk | -0.002 | -0.062 | 0.060 | 0.058 | 0.062 | TRUE |
| MRI_LeftPrecentral_vol | 0.013 | -0.003 | 0.016 | 0.014 | 0.017 | TRUE |
| MRI_LeftPrecuneus_thk | -0.004 | -0.002 | 0.001 | 0.000 | 0.003 | TRUE |
| MRI_LeftPrecuneus_vol | -0.007 | 0.008 | 0.015 | 0.014 | 0.017 | TRUE |
| MRI_LeftRostralanteriorcingulate_thk | -0.008 | -0.024 | 0.016 | 0.014 | 0.017 | TRUE |
| MRI_LeftRostralanteriorcingulate_vol | 0.002 | -0.023 | 0.025 | 0.023 | 0.026 | TRUE |
| MRI_LeftRostralmiddlefrontal_thk | 0.001 | -0.004 | 0.006 | 0.005 | 0.007 | TRUE |
| MRI_LeftRostralmiddlefrontal_vol | 0.008 | 0.020 | 0.012 | 0.011 | 0.014 | TRUE |
| MRI_LeftSuperiorfrontal_thk | 0.001 | -0.009 | 0.010 | 0.009 | 0.011 | TRUE |
| MRI_LeftSuperiorfrontal_vol | -0.006 | -0.016 | 0.010 | 0.009 | 0.012 | TRUE |
| MRI_LeftSuperiorparietal_thk | -0.003 | 0.000 | 0.003 | 0.002 | 0.004 | TRUE |
| MRI_LeftSuperiorparietal_vol | -0.036 | -0.006 | 0.030 | 0.028 | 0.032 | TRUE |
| MRI_LeftSuperiortemporal_thk | 0.006 | 0.008 | 0.002 | 0.001 | 0.003 | TRUE |
| MRI_LeftSuperiortemporal_vol | -0.002 | -0.018 | 0.017 | 0.015 | 0.018 | TRUE |
| MRI_LeftSupramarginal_thk | -0.037 | -0.044 | 0.007 | 0.004 | 0.009 | TRUE |
| MRI_LeftSupramarginal_vol | -0.030 | -0.001 | 0.029 | 0.027 | 0.030 | TRUE |
| MRI_LeftTemporalpole_thk | 0.021 | 0.006 | 0.015 | 0.014 | 0.017 | TRUE |
| MRI_LeftTemporalpole_vol | 0.009 | 0.024 | 0.015 | 0.014 | 0.017 | TRUE |
| MRI_LeftTransversetemporal_thk | 0.011 | 0.008 | 0.003 | 0.002 | 0.005 | TRUE |
| MRI_LeftTransversetemporal_vol | -0.007 | -0.045 | 0.039 | 0.037 | 0.040 | TRUE |
| MRI_RightBankssts_thk | -0.008 | 0.051 | 0.059 | 0.057 | 0.061 | TRUE |
| MRI_RightBankssts_vol | 0.001 | -0.048 | 0.049 | 0.047 | 0.050 | TRUE |
| MRI_RightCaudalanteriorcingulate_thk | 0.002 | 0.001 | 0.001 | 0.000 | 0.002 | FALSE |
| MRI_RightCaudalanteriorcingulate_vol | -0.017 | -0.046 | 0.029 | 0.027 | 0.031 | TRUE |
| MRI_RightCaudalmiddlefrontal_thk | -0.012 | 0.018 | 0.030 | 0.028 | 0.032 | TRUE |
| MRI_RightCaudalmiddlefrontal_vol | 0.019 | -0.023 | 0.042 | 0.040 | 0.044 | TRUE |
| MRI_RightCuneus_thk | 0.009 | 0.036 | 0.028 | 0.026 | 0.030 | TRUE |
| MRI_RightCuneus_vol | -0.009 | 0.003 | 0.011 | 0.010 | 0.013 | TRUE |
| MRI_RightEntorhinal_thk | 0.000 | 0.008 | 0.008 | 0.006 | 0.009 | TRUE |
| MRI_RightEntorhinal_vol | 0.005 | 0.032 | 0.027 | 0.025 | 0.028 | TRUE |
| MRI_RightFrontalpole_thk | 0.001 | 0.010 | 0.008 | 0.007 | 0.010 | TRUE |
| MRI_RightFrontalpole_vol | 0.006 | 0.000 | 0.006 | 0.005 | 0.007 | TRUE |
| MRI_RightFusiform_thk | -0.005 | -0.018 | 0.013 | 0.012 | 0.015 | TRUE |
| MRI_RightFusiform_vol | -0.003 | 0.012 | 0.015 | 0.014 | 0.017 | TRUE |
| MRI_RightHippocampus_vol | 0.000 | -0.037 | 0.036 | 0.034 | 0.038 | TRUE |
| MRI_RightInferiorparietal_thk | -0.013 | -0.008 | 0.005 | 0.003 | 0.006 | TRUE |
| MRI_RightInferiorparietal_vol | -0.003 | -0.044 | 0.041 | 0.040 | 0.043 | TRUE |
| MRI_RightInferiortemporal_thk | -0.001 | -0.004 | 0.003 | 0.002 | 0.004 | TRUE |
| MRI_RightInferiortemporal_vol | -0.056 | 0.021 | 0.077 | 0.075 | 0.079 | TRUE |
| MRI_RightInsula_thk | 0.025 | -0.009 | 0.034 | 0.033 | 0.036 | TRUE |
| MRI_RightInsula_vol | -0.017 | 0.036 | 0.053 | 0.051 | 0.055 | TRUE |
| MRI_RightIsthmuscingulate_thk | -0.045 | 0.001 | 0.046 | 0.044 | 0.047 | TRUE |
| MRI_RightIsthmuscingulate_vol | 0.013 | 0.001 | 0.012 | 0.011 | 0.013 | TRUE |
| MRI_RightLateraloccipital_thk | -0.008 | -0.004 | 0.005 | 0.003 | 0.006 | TRUE |
| MRI_RightLateraloccipital_vol | 0.002 | -0.041 | 0.043 | 0.041 | 0.044 | TRUE |
| MRI_RightLateralorbitofrontal_thk | -0.001 | -0.016 | 0.015 | 0.014 | 0.017 | TRUE |
| MRI_RightLateralorbitofrontal_vol | 0.003 | 0.024 | 0.021 | 0.019 | 0.022 | TRUE |
| MRI_RightLingual_thk | -0.006 | 0.055 | 0.061 | 0.059 | 0.063 | TRUE |
| MRI_RightLingual_vol | -0.007 | 0.004 | 0.011 | 0.010 | 0.012 | TRUE |
| MRI_RightMedialorbitofrontal_thk | 0.007 | 0.011 | 0.004 | 0.003 | 0.005 | TRUE |
| MRI_RightMedialorbitofrontal_vol | -0.004 | -0.014 | 0.010 | 0.008 | 0.011 | TRUE |
| MRI_RightMiddletemporal_thk | -0.001 | -0.022 | 0.021 | 0.020 | 0.023 | TRUE |
| MRI_RightMiddletemporal_vol | -0.001 | -0.083 | 0.082 | 0.079 | 0.084 | TRUE |
| MRI_RightParacentral_thk | -0.004 | 0.013 | 0.017 | 0.015 | 0.018 | TRUE |
| MRI_RightParacentral_vol | 0.003 | 0.016 | 0.013 | 0.012 | 0.014 | TRUE |
| MRI_RightParahippocampal_thk | -0.015 | -0.026 | 0.011 | 0.010 | 0.013 | TRUE |
| MRI_RightParahippocampal_vol | -0.007 | 0.010 | 0.017 | 0.016 | 0.018 | TRUE |
| MRI_RightParsopercularis_thk | 0.029 | 0.014 | 0.014 | 0.013 | 0.016 | TRUE |
| MRI_RightParsopercularis_vol | -0.002 | 0.028 | 0.030 | 0.029 | 0.032 | TRUE |
| MRI_RightParsorbitalis_thk | 0.026 | 0.016 | 0.010 | 0.009 | 0.012 | TRUE |
| MRI_RightParsorbitalis_vol | 0.017 | 0.011 | 0.006 | 0.004 | 0.007 | TRUE |
| MRI_RightParstriangularis_thk | 0.000 | -0.001 | 0.002 | 0.001 | 0.003 | TRUE |
| MRI_RightParstriangularis_vol | 0.017 | -0.046 | 0.062 | 0.060 | 0.064 | TRUE |
| MRI_RightPericalcarine_thk | 0.000 | -0.003 | 0.003 | 0.002 | 0.004 | TRUE |
| MRI_RightPericalcarine_vol | -0.013 | -0.004 | 0.008 | 0.007 | 0.010 | TRUE |
| MRI_RightPostcentral_thk | -0.002 | 0.004 | 0.007 | 0.005 | 0.008 | TRUE |
| MRI_RightPostcentral_vol | -0.002 | 0.045 | 0.047 | 0.046 | 0.049 | TRUE |
| MRI_RightPosteriorcingulate_thk | 0.005 | 0.011 | 0.006 | 0.005 | 0.007 | TRUE |
| MRI_RightPosteriorcingulate_vol | 0.001 | 0.017 | 0.016 | 0.014 | 0.017 | TRUE |
| MRI_RightPrecentral_thk | -0.002 | -0.021 | 0.020 | 0.018 | 0.021 | TRUE |
| MRI_RightPrecentral_vol | 0.004 | 0.019 | 0.015 | 0.013 | 0.016 | TRUE |
| MRI_RightPrecuneus_thk | 0.000 | -0.013 | 0.013 | 0.012 | 0.015 | TRUE |
| MRI_RightPrecuneus_vol | 0.009 | 0.001 | 0.008 | 0.007 | 0.010 | TRUE |
| MRI_RightRostralanteriorcingulate_thk | 0.001 | -0.012 | 0.013 | 0.012 | 0.014 | TRUE |
| MRI_RightRostralanteriorcingulate_vol | -0.014 | 0.056 | 0.070 | 0.068 | 0.072 | TRUE |
| MRI_RightRostralmiddlefrontal_thk | 0.006 | 0.002 | 0.004 | 0.003 | 0.006 | TRUE |
| MRI_RightRostralmiddlefrontal_vol | -0.005 | 0.025 | 0.031 | 0.029 | 0.032 | TRUE |
| MRI_RightSuperiorfrontal_thk | -0.002 | 0.021 | 0.023 | 0.022 | 0.025 | TRUE |
| MRI_RightSuperiorfrontal_vol | 0.001 | -0.019 | 0.020 | 0.018 | 0.021 | TRUE |
| MRI_RightSuperiorparietal_thk | -0.022 | -0.031 | 0.009 | 0.007 | 0.011 | TRUE |
| MRI_RightSuperiorparietal_vol | 0.008 | 0.023 | 0.015 | 0.013 | 0.017 | TRUE |
| MRI_RightSuperiortemporal_thk | -0.006 | 0.036 | 0.042 | 0.040 | 0.045 | TRUE |
| MRI_RightSuperiortemporal_vol | -0.004 | -0.024 | 0.021 | 0.019 | 0.023 | TRUE |
| MRI_RightSupramarginal_thk | 0.009 | -0.009 | 0.018 | 0.016 | 0.019 | TRUE |
| MRI_RightSupramarginal_vol | -0.003 | 0.009 | 0.012 | 0.011 | 0.013 | TRUE |
| MRI_RightTemporalpole_thk | 0.003 | -0.004 | 0.007 | 0.005 | 0.008 | TRUE |
| MRI_RightTemporalpole_vol | -0.018 | 0.021 | 0.039 | 0.038 | 0.041 | TRUE |
| MRI_RightTransversetemporal_thk | -0.006 | 0.028 | 0.034 | 0.033 | 0.036 | TRUE |
| MRI_RightTransversetemporal_vol | -0.001 | -0.003 | 0.001 | 0.000 | 0.002 | TRUE |
| MRI_TotalGrayMatter_vol | -0.002 | 0.013 | 0.015 | 0.013 | 0.016 | TRUE |
| Pulse | 0.007 | -0.003 | 0.010 | 0.009 | 0.011 | TRUE |
| SmokeYN_1.0 | 0.005 | -0.028 | 0.033 | 0.030 | 0.035 | TRUE |
| TAU_Age | -0.140 | -0.090 | 0.051 | 0.048 | 0.054 | TRUE |
| Weight | -0.010 | 0.003 | 0.013 | 0.011 | 0.014 | TRUE |
| YrsEd | -0.001 | 0.021 | 0.023 | 0.021 | 0.024 | TRUE |
| lPACC | -0.078 | -0.153 | 0.074 | 0.072 | 0.077 | TRUE |
